# Identifying the neural correlates of anticipatory postural control: a novel fMRI paradigm

**DOI:** 10.1101/2022.09.25.22280328

**Authors:** Jo Armour Smith, Rongwen Tain, Kelli G. Sharp, Laura M. Glynn, Linda R. Van Dillen, Korinne Henslee, Jesse V. Jacobs, Steven C. Cramer

## Abstract

**Introduction:** Postural control is essential for maintaining body equilibrium during voluntary limb movement. Altered postural control in the trunk and hip musculature is a characteristic of aging and of multiple neurological and musculoskeletal conditions. Due to the difficulty of designing a task for the MRI environment that elicits postural activation in the trunk/hip musculature, it has not previously been possible to determine if altered cortical and subcortical sensorimotor brain activation underlies observed impairments in postural control in patient populations. The purpose of this study was to use a novel fMRI-compatible paradigm to identify the sensorimotor brain activation associated with anticipatory postural control in the trunk and hip musculature in healthy adults.

**Methods:** BOLD fMRI imaging was performed on 20 healthy volunteers (23 ± 4 years, 13 female, 7 male, Siemens Prisma 3T MRI). Participants performed two versions of a lower limb task involving lifting the left leg a short distance to touch the foot to a horizontal target. For the supported leg raise task (SLR) the leg is raised from the knee while the thigh remains supported. For the unsupported leg raise task (ULR) the leg is raised from the hip. Anticipatory postural muscle activation is elicited in the bilateral abdominal and contralateral hip extensor musculature during the ULR but not the SLR. Thirty-two repetitions were completed for each task in response to visual cues using an event-related design. Data were processed using SPM12 and framewise head displacement was quantified using the Artifact Detection Tool. Anatomical masks for primary and secondary sensory and motor cortical regions and for the cerebellum and basal ganglia were created using WFU-PickAtlas for the right and left sides separately.

**Results:** Framewise head displacement was within acceptable limits for both tasks (SLR 0.27 ± 0.1mm, ULR 0.18 ± 0.1 mm). Significant brain activation during the SLR task occurred predominantly in the right primary and secondary sensorimotor cortical regions. Brain activation during the ULR task occurred bilaterally in the primary and secondary sensorimotor cortical regions, as well as cerebellum and putamen. In comparison with the SLR, the ULR was associated with significantly greater activation in the right premotor/SMA, left primary motor and cingulate cortices, primary somatosensory cortex, supramarginal gyrus/parietal operculum, superior parietal lobule, cerebellar vermis, and bilateral cerebellar hemispheres.

**Conclusions:** Cortical and subcortical regions activated during the unsupported leg raise, but not during the supported leg raise, were consistent with the planning, execution, and sensory experience of a task involving multi-segmental and bilateral postural control. This paradigm provides a foundation for future studies that will isolate neural mechanisms underlying impaired postural control in patients with neurological and musculoskeletal dysfunction.

## INTRODUCTION

Postural control is an essential component of many voluntary movements. When performing a motor task such as reaching to grasp an object or shifting weight to take a step, synergies of postural activation in the trunk and hip musculature help to maintain the upright orientation of body segments and to preserve balance during the focal movement. The control mechanisms that underlie postural synergies include both feedforward and feedback processes. Feedforward postural muscle activation occurs immediately prior to or at the same time as the initiation of an anticipated voluntary movement. This feedforward activation mitigates the effects of the perturbing forces associated with voluntary movement and is termed an anticipatory postural adjustment (APA).^1^

Altered postural control of the trunk and hip musculature is a characteristic of aging and of multiple neurological and musculoskeletal conditions.^2–7^ Impaired APAs result in greater reliance on feedback mechanisms of postural control, dysfunctional joint loading, and reduced ability to maintain balance during dynamic movement.^8–10^ Human studies using electroencephalography (EEG), non-invasive brain stimulation, or investigation of individuals with brain lesions have identified multiple sensorimotor cortical regions that appear to contribute to anticipatory postural control. These include the primary motor cortex, the lateral premotor area, and the supplementary motor area (SMA).^11–14^ In contrast, the sub-cortical neural correlates of APAs have been more difficult to determine. Impairment of APAs in individuals with Parkinson’s disease suggests that the basal ganglia are involved in anticipatory postural control.^15,16^ Studies investigating the involvement of the cerebellum in production of APAs using healthy individuals and patient populations have been inconclusive.^17–20^ One paradigm has used magnetoencephalography during a supine bimanual task to explore sub-cortical postural planning.^20^ However, upper limb movements conducted in supine are unlikely to elicit significant trunk or hip activation and so may not generalize to the postural control associated with lower limb motion.^21^

To understand how functional brain reorganization may contribute to impaired postural control in individuals with neurological or musculoskeletal dysfunction, it is critical to first determine the neural correlates of postural control in healthy individuals. Two recent preliminary studies described an fMRI-compatible lower limb paradigm that enables measurement of the cortical and sub-cortical activation associated with anticipatory postural control in the trunk and hip.^4,22^ This paradigm involves two small-amplitude leg movement tasks. One task, the supported leg raise, does not require postural activation of the trunk/hip. In the other task, the unsupported leg raise, anticipatory postural muscle activation is elicited in the bilateral abdominal and contralateral hip extensor musculature.^22,23^ The preliminary studies demonstrated the feasibility of the paradigm in a single participant^22^ and established an association between the fMRI paradigm and postural control demonstrated outside of the scanner environment.^24^ The purpose of this study was to use the novel fMRI-compatible paradigm to identify the sensorimotor brain activation associated with anticipatory postural control in the trunk and hip musculature in healthy adults.

## MATERIALS AND METHODS

### Participants

Twenty healthy volunteers (13 female, age 23 ± 4 years, body mass index 21.6 kg/m^2^) participated in the study. Sample size for adequate statistical power was calculated based on existing literature.^25,26^ Participants were eligible for inclusion if they reported being right-handed and were between the ages of 19 and 35 years. Exclusion criteria included history of back pain or other chronic pain condition requiring medical care or resulting in limitation of function, history of inflammatory or neurological disorders, and any contraindication to MRI scanning. The study was approved by the Institutional Review Board at Chapman University, and participants gave written informed consent prior to participating.

### Experimental procedure

Limb preference for both the upper limb and lower limb was quantified using the Lateral Preference Inventory (LPI, 4-item handedness and footedness subscales).^27^ A score of 4 indicates consistent right limb preference and -4 indicates consistent left limb preference.

Scanning was conducted using a Siemens MAGNETOM 3T Prisma scanner (Siemens Medical Solutions USA Inc, PA, USA) and a 32-channel head coil. Head stabilization was provided by padding around the neck and head and by a chin strap. Participants performed the two lower limb tasks. Both tasks involved lifting the leg a short distance until they felt the ankle touch a horizontal target. The height of the target above the support surface was individualized and set at half of the length of the participant’s shank (distance from tibial tuberosity to base of calcaneus, approximately 20 cm, on average). For the supported leg raise (SLR) task, participants were positioned in hip and knee flexion with a 14 cm wedge under the knees and the arms by their sides. The lower leg was raised to touch the foot to the target by extending the knee while the knee and thigh remained supported (Figure 1: A). For the unsupported leg raise (ULR) task, the participant was positioned with the hip and knee extended and their arms by their sides. The entire leg was raised to touch the foot to the target by flexing the hip (Figure 1: A).^4,22^ Participants had extensive practice of the ULR and SLR on a separate study visit prior to the imaging visit, and all performed the task with their left limb.

**Figure 1.**
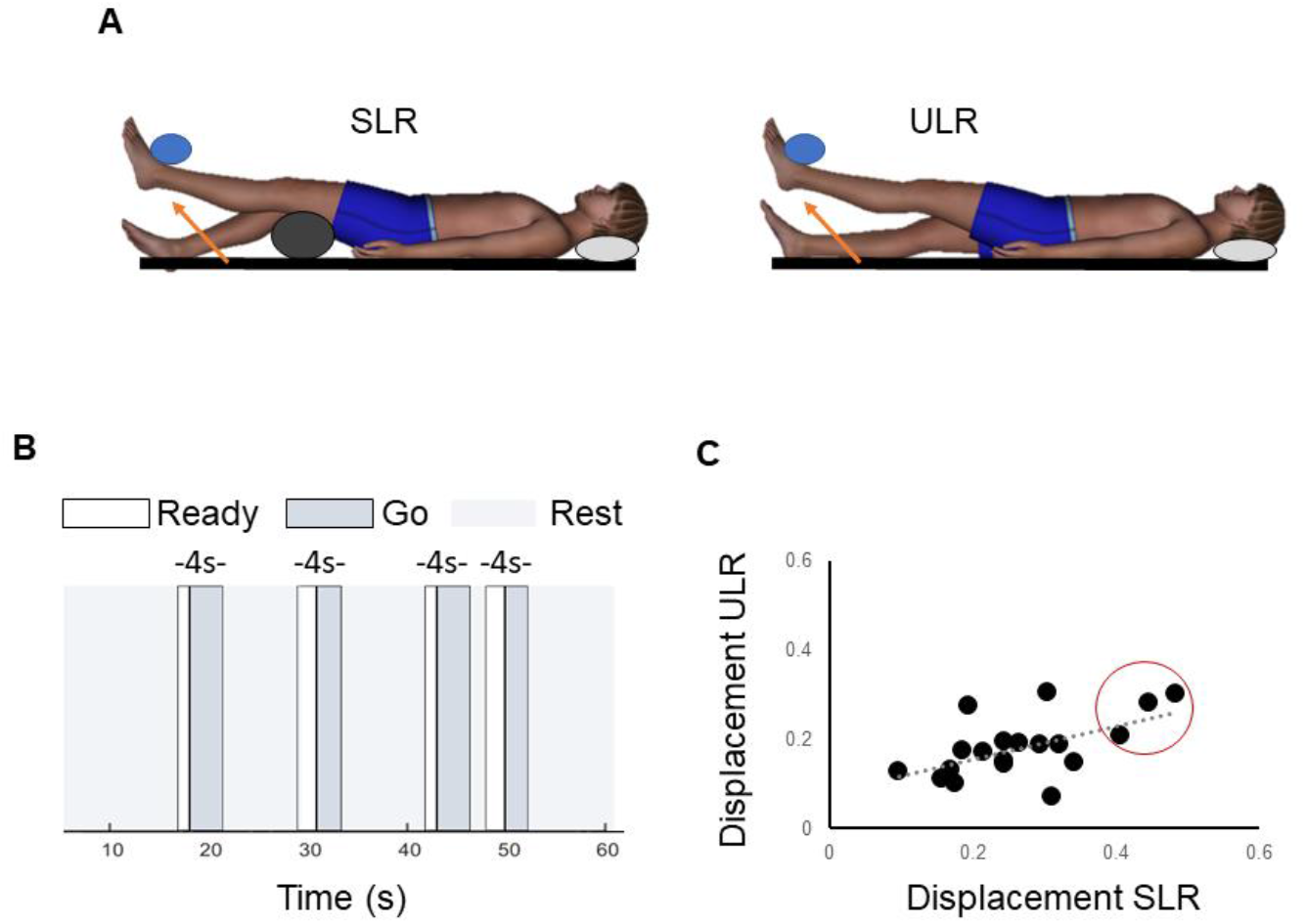
A) Schematic showing supported leg raise (SLR) and unsupported leg raise (ULR). B) Example of stimuli during event-related paradigm. C) Scatter plot showing relationship between mean framewise mm of head displacement during the supported leg raise (SLR) and unsupported leg raise (ULR) in the twenty participants. Individuals circled in red were excluded from the SLR analysis and task comparisons due to exceeding the a priori threshold for head displacement.

An event-related paradigm was used to quantify sensorimotor activation. Participants performed one run of each task, with 32 repetitions of the leg raise and 32 relaxation periods in each run. The following visual stimuli were provided for the initiation and end of each leg raise repetition: “Ready”, “Go”, and “Rest”. Participants were instructed to get ready to move when the “Ready” stimulus was displayed, to slowly perform the leg raise to touch the foot to the target when the “Go” stimulus was displayed, and to slowly lower the leg back to the starting position when the “Rest” stimulus was displayed. The duration of the “Ready” stimulus varied randomly from 1 to 2 seconds (in 0.5 s increments). The duration of the “Go” stimulus varied from 2 to 3 seconds (in 0.5 s increments). The total duration of the “Ready” and “Go” stimuli was 4 seconds for all repetitions (Figure 1:B). The 32 relaxation (no movement) periods each had a duration of 4 seconds and were interspersed between movement repetitions. During the relaxation periods the “Rest” stimulus remained visible to the participants. The ordering of leg raise and relaxation events and the inter-stimulus intervals were optimized and jittered to maximize the hemodynamic response function for the contrast of leg raise versus relaxation using the OptSeq2 event scheduling tool.^28,29^ The best two sequences generated by OptSeq2 were used, counterbalanced across participants.

The visual stimuli were synched with the scanner and presented to the participants using PsychoPy.^30,31^ Log files with the stimulus onset times for each run were saved and utilized in the first-level models. Participant performance was monitored by the investigator throughout the scanning period. At the end of each task participants were asked to quantify any pain or discomfort during the task on a 0 to 10 numeric rating scale.

### Image acquisition

T2*-weighted echo planar images with blood oxygen level-dependent (BOLD) contrast were acquired with the following parameters: repetition time (TR) 2000 ms, time to echo (TE) 34.5 ms, voxel size 3 × 3 × 3 mm^3^, flip angle 70º, 56 slices, scan time 418 seconds, 209 volumes per run. Field-map images were acquired prior to each task run. In addition, an anatomical T1-weighted scan was acquired at the beginning of the session (MPRAGE, TR 2400 ms, TE 2.3 ms, voxel size 0.7 × 0.7 × 0.7 mm^3^).

Functional MRI data were pre-processed using SPM12 (v7771, The Wellcome Centre for Human Neuroimaging, London, UK), running in Matlab R2018b (Mathworks, MA, USA). Images were inspected and manually reoriented as needed. Realignment translation and rotation parameters were calculated using the INRIalign toolbox.^32^ Images were unwarped to correct for B0 inhomogeneities and slice time corrected.^33^ The anatomical scans were co-registered with the mean functional images using the normalized mutual information approach and were normalized to MNI space using unified segmentation.^34^ The functional images were then spatially normalized using 4^th^ degree B-spline interpolation, resliced to 2 mm isotropic voxel size, and smoothed with an 8 mm full width at half maximum (FWHM) Gaussian kernel.

Framewise head displacement was quantified using the Artifact Detection Tool (www.nitrc.org/projects/artifact_detect/).^35^ Total framewise displacement was calculated as the root mean square of all three directions of translation plus each rotation (multiplied by 65 to convert rotations into absolute distance) in mm.^36^ Volumes with greater than 0.5 mm/TR motion^37,38^ were repaired using linear interpolation of values from adjacent volumes.^35^ The threshold for exclusion of participants was set *a priori* as those with greater than 30% of volumes requiring repair.^39,40^

For the first-level analysis (individual participant level), the timing of the leg raise (starting at the “Ready” stimulus) and the relaxation events was convolved with the hemodynamic response function. The duration of each predictor event was modeled as 4 seconds. Individual t-contrasts were calculated for SLR > relax and ULR > relax using a general linear model for each participant. The first 9 volumes of each scan were discarded to ensure equilibrium of the signal, leaving 30 leg raises and relaxation events for analysis in each task. The realignment parameters were entered as regressors of no interest and the data were high-pass filtered at 128 Hz.

For the second-level analysis (group level), the individual t-contrasts were entered into random effects models to test the contrasts SLR > relax and ULR > relax. Initially, whole brain analyses were conducted with a family-wise error (FWE) corrected threshold of p < 0.05. Then, anatomical masks were created using WFU-PickAtlas^10^ for the right and left sides separately. Regions of interest (ROI) were determined *a priori* based on existing literature and comprised the following areas: primary motor cortex, premotor cortex/supplementary motor area (SMA), midcingulate cortex, primary somatosensory cortex, superior parietal lobule, supramarginal gyrus/parietal operculum, putamen, globus pallidus, caudate, cerebellar vermis (single midline ROI), and the cerebellar hemispheres (motor areas).^25,38,41–43^

The magnitude of activation within each ROI was assessed by calculating peak percent signal change in the ROI for each individual, scaled by normalization to each individual’s mean baseline activation, by the peak regressor within the design matrix, and by the contrast sum.^44,45^ After confirming that data met assumptions of normality and variance, separate two-way ANOVAs were used to test for main effects of task, ROI, and task by ROI interactions for the right hemisphere cortical ROIs, left hemisphere cortical ROIs, and the sub-cortical ROIs. When significant task or task*ROI interactions were observed, post-hoc comparisons between tasks were completed using paired t-tests with Holm-Bonferroni correction for multiple comparisons. To assess the lateralization of activation in the cortical ROIs during each task, a laterality index was calculated from the signal change data using the equation LI = (C – I)/(C + I) such that C is the magnitude of activation in the hemisphere contralateral to the leg raise (right) and I is the magnitude of activation in the hemisphere ipsilateral to the leg raise (left).^46,47^ Extent of activation within each ROI was assessed by calculating the proportion of voxels within the ROI that were activated at the FWE-corrected threshold of p < 0.05. Differences in extent of activation between tasks were compared using two-way ANOVA as described above.

Finally, an exploratory whole-brain analysis was conducted using a direct subtraction approach with the contrast ULR > SLR at the first and second levels and a cluster-level FWE corrected threshold of p < 0.05.

## RESULTS

No participants reported any pain or discomfort during the leg raise tasks. Median LPI score was 4 for both handedness and footedness, indicating consistent right hand and right foot preference.^27^ One participant scored 0 for handedness, and two participants scored 0 for footedness, indicating ambilaterality.

Mean framewise displacement did not exceed 0.5 mm in any individual for either task. Across the group, head displacement was significantly greater during the supported leg raise than the unsupported leg raise (SLR 0.27 ± 0.1 mm, ULR 0.18 ± 0.1 mm, p < 0.001). Individuals with greater head displacement in the supported leg raise task also had greater head displacement in the unsupported task (r = 0.565, p = 0.012, Figure 1:C). For the SLR, there was no correlation between extent of head motion and peak brain activation in any of the cortical or sub-cortical ROIs except the right cerebellar hemisphere where a negative relationship was observed (r = -0.555, p = 0.021). For the ULR there was no correlation between extent of head motion and peak brain activation in any of the ROIs. Head motion during both tasks did not differ by sex (SLR p = 0.194, ULR p = 0.351) but was significantly correlated with body weight (SLR r = 0.633, p = 0.003, ULR r = 0.565, p = 0.012).

Three participants (one female and two males, identified by the red circle in Figure 1: C) exceeded the threshold determined *a priori* for number of volumes requiring repair for the SLR. They were excluded from the SLR analyses and task comparisons presented here. However, a sensitivity analysis indicated that results did not differ if all 20 participants were included in the SLR analysis or if the same 3 participants were excluded from the ULR analysis. Average percentage of repaired volumes in the remaining participants was 10% (± 10%) for the SLR and 4% (± 6%) for the ULR.

### Supported leg raise – whole brain analysis

During the supported leg raise, significant activation occurred in the right paracentral lobule (encompassing both the primary motor and sensory cortices), and in the right midcingulate cortex (Figure 2: A and Table 1). There was also significant activation bilaterally in the supramarginal gyri, the supplementary motor areas, and in the midline in the cerebellar vermis.

**Figure 2.**
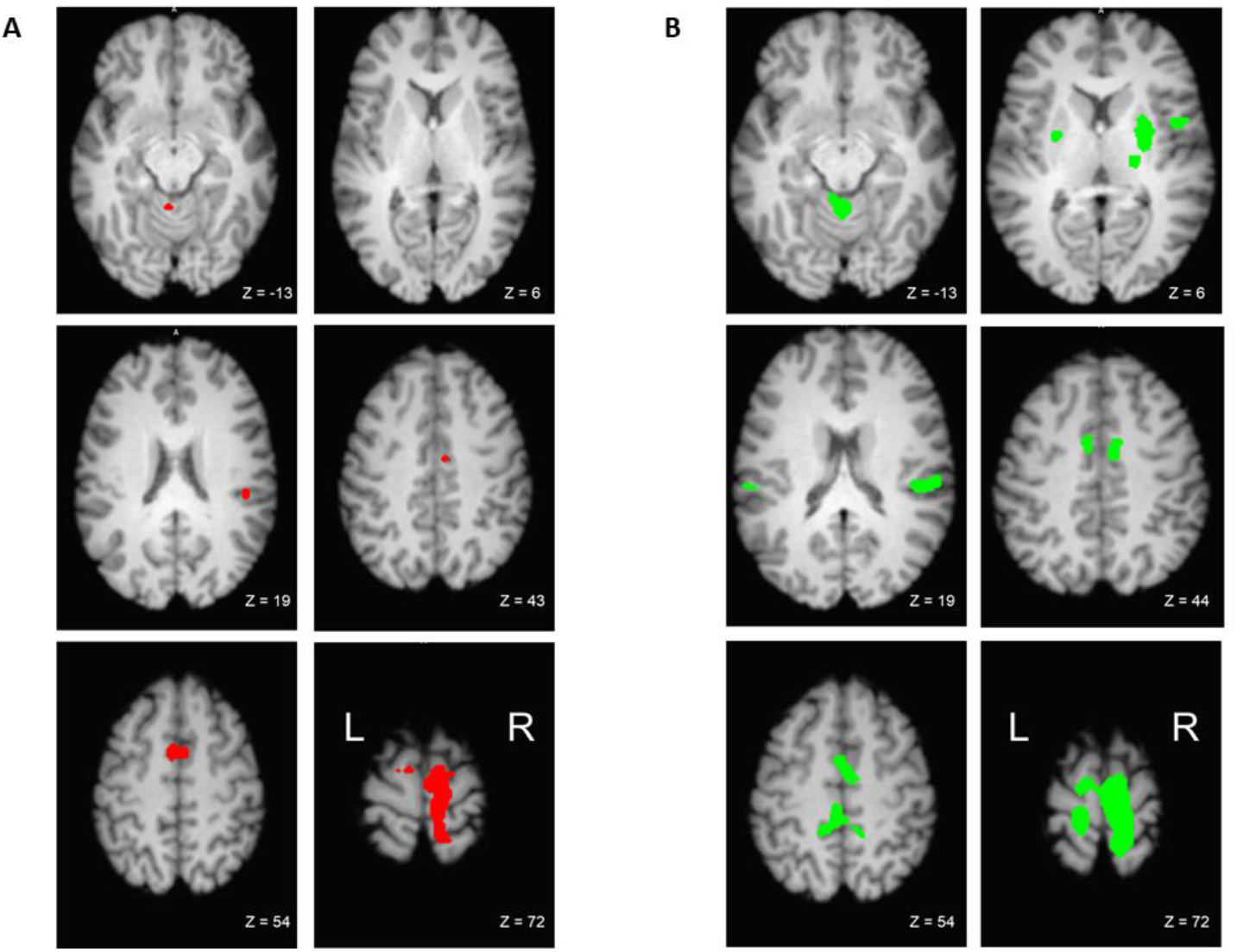
Significant brain activation during A) supported leg raise and B) unsupported leg raise. FWE corrected at p < 0.05.

**Table 1.** Significant whole brain activation for the contrast supported leg raise > relax

| Location | BA | Cluster size | Percent signal change | Z | MNI coordinates |  |  |
| --- | --- | --- | --- | --- | --- | --- | --- |
|  |  |  |  |  | x | y | z |
| Right paracentral lobule | 4 | 1032 | 0.56 (0.49 – 0.62) | 6.46 | 8 | -28 | 64 |
| Right supplementary motor area |  |  | 1.68 (1.47 – 1.90) | 6.16 | 8 | -12 | 76 |
| Right supplementary motor area | 6 |  | 0.69 (0.60 – 0.78) | 6.15 | 8 | -14 | 68 |
| Left supplementary motor area | 6 | 81 | 0.53 (0.45 – 0.60) | 5.90 | -4 | 4 | 52 |
| Right supplementary motor area | 6 |  | 0.39 (0.31 – 0.47) | 4.97 | 8 | 0 | 52 |
| Right supramarginal gyrus |  | 20 | 0.54 (0.46 – 0.63) | 5.62 | 46 | -28 | 24 |
| Right midcingulate | 24 | 6 | 0.37 (0.31 – 0.48) | 5.46 | 8 | -10 | 42 |
| Lobule IV, V of vermis |  | 9 | 0.46 (0.37 – 0.54) | 5.18 | -2 | -48 | -14 |
| Left supramarginal gyrus | 40 | 5 | 0.55 (0.47 – 0.64) | 5.14 | -50 | -40 | 24 |

Threshold $p < 0.05$ FWE-corrected. Minimum cluster size of 5 voxels, reported in MNI space. Cluster size is in number of voxels (voxel dimensions $3\text{mm}^3$ ). Locations of significant local maxima are listed and labeled using the Automated Anatomical Labelling Atlas (AAL). Where applicable cytoarchitectonic areas (Brodmann areas, BA) are also provided and defined using the SPM Anatomy Toolbox and the Bioimage MNI2Tal tool. Mean percent signal change (90% confidence interval) calculated using individual scaling factors.

### Unsupported leg raise – whole brain analysis

During the unsupported leg raise, a large bilateral area of activation encompassed the primary motor and sensory cortices, the supplementary motor areas, and the left midcingulate cortex. Bilateral activation also occurred in the supramarginal gyri, in the putamen and in lobules IV and V of the cerebellar hemispheres. In the right hemisphere, there was additional activation in the thalamus and the Rolandic operculum (Figure 2: B and Table 2).

**Table 2.** Significant whole brain activation for the contrast unsupported leg raise > relax

| Location | BA | Cluster size | Percent signal change | Z | MNI coordinates |  |  |
| --- | --- | --- | --- | --- | --- | --- | --- |
|  |  |  |  |  | x | y | z |
| Right postcentral gyrus | 1 | 2876 | 0.91 (0.82 – 1.00) | 7.11 | 12 | -34 | 68 |
| Right precuneus |  |  | 1.07 (0.95 – 1.19) | 6.87 | 12 | -44 | 64 |
| Right supplementary motor area | 6 |  | 0.80 (0.62 – 0.80) | 6.61 | 12 | -16 | 70 |
| Right putamen |  | 157 | 0.30 (0.24 – 0.41) | 5.81 | 28 | -4 | 8 |
| Right putamen |  |  | 0.27 (0.21 – 0.32) | 5.31 | 26 | 6 | 8 |
| Right thalamus |  | 46 | 0.30 (0.25 – 0.35) | 5.72 | 20 | -20 | 10 |
| Right rolandic operculum | 40 | 100 | 0.60 (0.50 – 0.71) | 5.70 | 46 | -26 | 20 |
| Right supramarginal gyrus | 40 |  | 0.58 (0.47 – 0.70) | 5.30 | 56 | -24 | 18 |
| Right midcingulate | 32 | 67 | 0.53 (0.44 – 0.63) | 5.62 | 8 | -4 | 42 |
| Lobule IV, V of vermis |  | 208 | 0.70 (0.58 – 0.83) | 5.61 | 0 | -50 | -12 |
| Lobule III of vermis |  |  | 0.61 (0.49 – 0.72) | 5.44 | -2 | -46 | -22 |
| Lobule IV, V of left cerebellar hemisphere |  |  | 0.59 (0.47 – 0.70) | 5.40 | -8 | -42 | -16 |
| Right rolandic operculum | 6 | 54 | 0.44 (0.36 – 0.52) | 5.51 | 48 | 4 | 8 |
| Lobule IV, V of left cerebellar hemisphere |  | 33 | 0.60 (0.48 – 0.72) | 5.36 | -18 | -38 | -24 |
| Left putamen |  | 18 | 0.24 (0.19 – 0.30) | 5.13 | -26 | -4 | 6 |
| Lobule IV, V of right cerebellar hemisphere |  | 9 | 0.36 (0.28 – 0.44) | 5.01 | 24 | -36 | -30 |
| Left supramarginal gyrus | 40 | 14 | 0.56 (0.42 – 0.68) | 4.95 | -58 | -26 | 20 |
| Lobule VIII of vermis |  | 7 | 0.49 (0.47 – 0.75) | 4.94 | 0 | -68 | -44 |

Threshold $p < 0.05$ FWE-corrected. Minimum cluster size of 5 voxels, reported in MNI space. Cluster size is in number of voxels (voxel dimensions 3mm<sup>3</sup>). Locations of significant local maxima are listed and labeled using the Automated Anatomical Labelling Atlas (AAL). Where applicable cytoarchitectonic areas (Brodmann areas, BA) are also provided and defined using the SPM Anatomy Toolbox and the Biomed MNI2Tal tool. Mean percent signal change (90% confidence interval) calculated using individual scaling factors.

### Task comparison - signal change

Percent signal change for ROIs in the right cortex during both tasks is shown in Figure 3: A. There were significant main effects of task (F = 5.983, p = 0.026) and ROI (F = 13.861, p < 0.001), and a task by ROI interaction (F = 2.604, p = 0.031). Holm-Bonferroni corrected post-hoc pairwise comparisons between tasks showed that there was significantly greater activation in the right premotor/SMA ROI during the ULR than the SLR (adjusted p = 0.024). Task-dependent activation differences in the cingulate and primary sensorimotor ROI did not survive adjustment for multiple comparisons.

**Figure 3.**
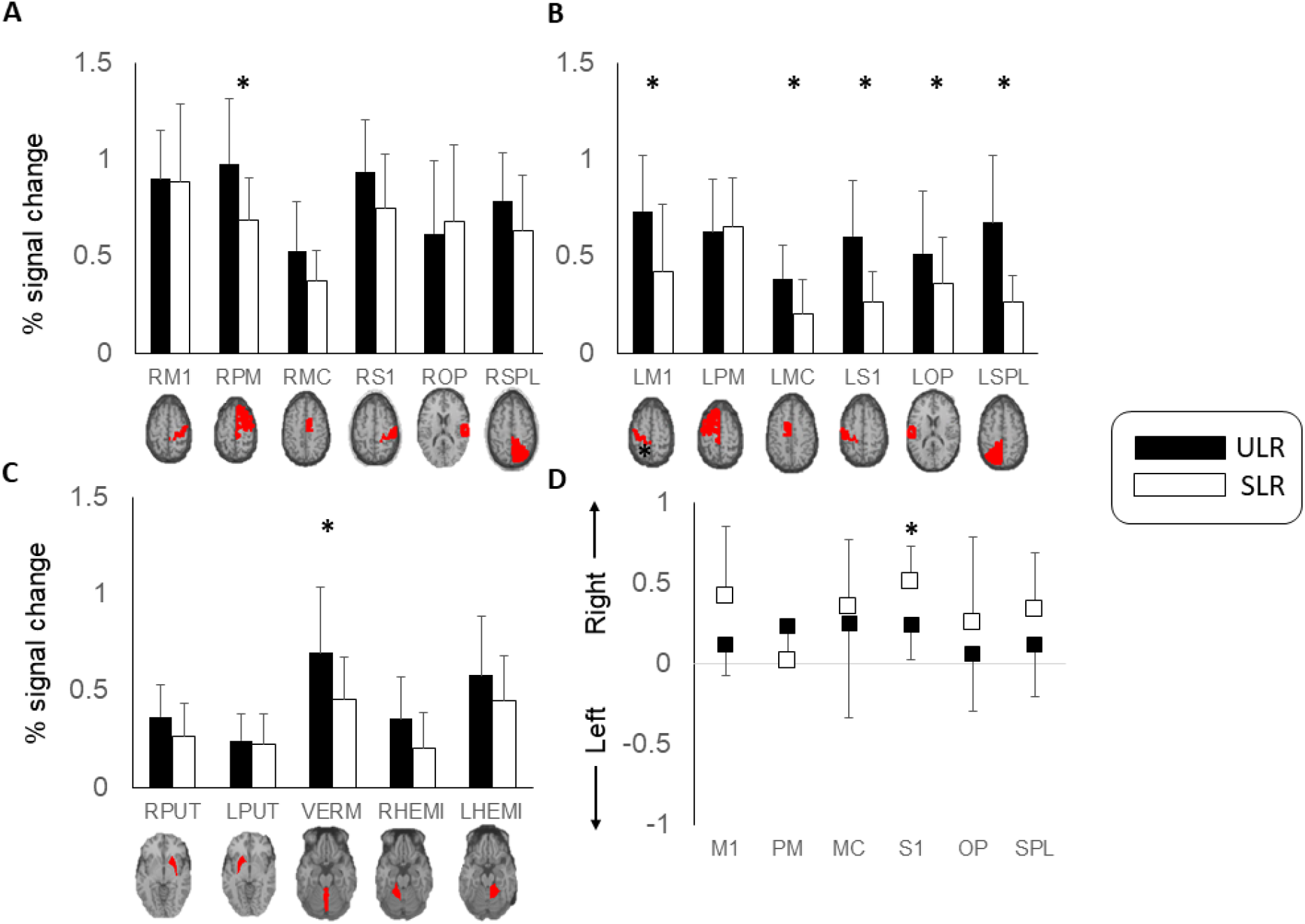
Mean ± standard deviation percent signal change in regions of interest. A) Right hemisphere. B) Left hemisphere. C) Sub-cortical. D) Lateralization index for cortical regions of interest. M1 – primary motor cortex. PM – premotor cortex/supplementary motor area. MC – midcingulate cortex. S1 – primary somatosensory cortex. OP – supramarginal gyrus/parietal operculum. SPL – superior parietal lobule. PUT – putamen. VERM – cerebellar vermis (single midline ROI). HEMI – cerebellar hemisphere.

Percent signal change for ROIs in left cortex during both tasks are shown in Figure 3: B. There were significant main effects of task (F = 30.154, p < 0.001) and ROI (F = 9.151, p < 0.001) and a task by ROI interaction (F = 12.273, p < 0.001). Holm-Bonferroni corrected post-hoc comparisons showed that there was significantly greater activation in the following ROIs during the ULR compared with the SLR: primary motor (p < 0.001), cingulate (p = 0.006), primary somatosensory (p < 0.001), supramarginal gyrus/parietal operculum (p = 0.032) and superior parietal lobule (p < 0.001).

Percent signal change for ROIs in right and left sub-cortical ROIs during both tasks are shown in Figure 3: C (caudate and globus pallidus ROIs not shown due to low levels of activity). On the right, there were significant main effects of task (F = 7.180, p = 0.016) and ROI (F = 14.000, p < 0.001). Differences between tasks were not significant following adjustment for multiple comparisons. On the left, there were significant main effects of ROI (F = 14.000, p < 0.001) but no main effect of task or task by ROI interaction. Activation in the cerebellar vermis was greater in the ULR task (p < 0.001).

### Task comparison - proportion of active voxels

The largest proportion of active voxels occurred in the right and left premotor ROIs for both tasks (right hemisphere; ULR 22.4 (± 12.2)%, SLR 15.4 (± 13.6)%, left hemisphere; ULR 16.3 (±10.8)%, SLR 8.9 (±11.2)%).

For ROIs in the right hemisphere there was no significant main effect of task (F = 3.570, p = 0.077) or task by ROI interaction (F = 0.797, p = 0.555). Proportion of activation did vary significantly by ROI (F = 22.041, p < 0.001). For ROIs in the left hemisphere there was a significant main effect of task (F = 4.641, p = 0.047) and ROI (F = 2.434, p = 0.027) but no task by ROI interaction (F = 2.434, p = 0.096). Holm-Bonferroni corrected post-hoc comparisons showed that there was significantly greater proportion of active voxels in primary motor cortex during the ULR (ULR; 13.7 (±10.9)%, SLR 6.1 (±8.5)%, p = 0.024). There was greater inter-individual variability in the proportion of voxels that exceeded the FWE-rate corrected threshold for significant activation in the basal ganglia ROI. Few participants had significantly activated voxels in the caudate nuclei or globus pallidus bilaterally. Fifteen participants demonstrated active voxels in the right putamen during the ULR, but only seven had activation in the same ROI for the SLR. There was more consistent activation in the cerebellar vermis and cerebellar hemispheres and task comparisons for these ROIs were conducted using Wilcoxon Signed Rank tests. The proportion of active voxels was significantly greater in all three cerebellar ROIs during the ULR (vermis ULR; 15.9 (±16.8)%, vermis SLR 3.7 (± 5.3)%, right cerebellar hemisphere ULR; 5.9 (± 8.7)%, right cerebellar hemisphere SLR; 2.1 (± 8.4)%, left cerebellar hemisphere ULR; 12.8 (±13.3)%, left cerebellar hemisphere SLR 4.8 (± 8.7)%, p < 0.02 for all comparisons).

### Task comparison - laterality index

Average laterality index for each ROI for each task is shown in Figure 3: D. As expected, greater laterality (activation contralateral to the moving limb) was evident during the SLR than the ULR. There were significant main effects of task (F = 9.157, p = 0.009) and ROI (F = 4.029, p = 0.003), and a task by ROI interaction (F = 3.079, p = 0.014). Holm-Bonferroni corrected post-hoc pairwise comparisons between tasks showed that there was significantly lower laterality index, indicating more activation ipsilateral to the moving limb, during the ULR in the primary somatosensory cortex (adjusted p = 0.006).

### Task comparison – direct subtraction ULR - SLR

For the subtraction analysis, areas of activation that were significantly greater during the ULR than the SLR are shown in Figure 4: A and listed in Figure 4: B. Significantly greater activation occurred in the left paracentral lobule (encompassing both the primary motor and sensory cortices), in the cerebellar vermis, and in the left cerebellar hemisphere.

**Figure 4.**
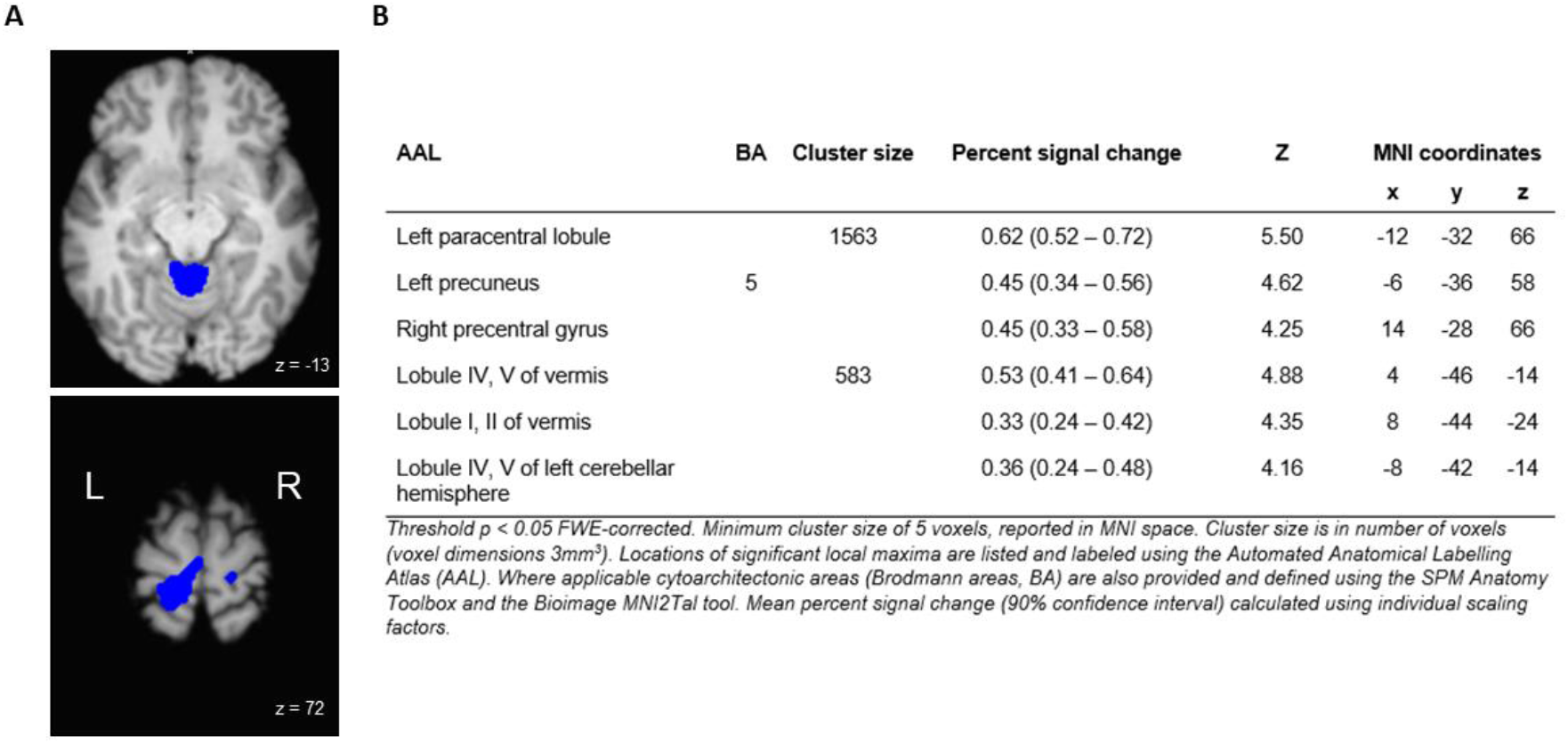
A) and B) Significant whole brain activation for the contrast unsupported leg raise > supported leg raise.

## DISCUSSION

We identify the cortical and sub-cortical sensorimotor brain activation associated with postural control in the trunk and hip musculature during voluntary lower limb movement. Impaired postural control in the trunk and hip is common, e.g., in individuals with musculoskeletal pain, neurological dysfunction, and in older adults.^2,3,5–7,48^ The fMRI-compatible leg raise paradigm tested in this study provides insights into control of APAs and will enable future research to establish cortical and sub-cortical contributors to altered postural control in patient populations.

Our research builds upon preliminary work that used this experimental approach but did not report magnitude or extent of activation in specific sensorimotor regions of interest.^4^ To our knowledge, this paradigm is the first to permit measurement of whole brain neural correlates of postural control during a lower limb movement. Recent work by Silfies et al.,^25^ and Jordan et al.,^26^ used a task involving unilateral and bilateral spinal and hip extension (bridging) to demonstrate sensorimotor control of movement in this region. With this bridging paradigm it is more difficult to separate activation associated with the focal and postural components of the movement. However, the percent signal change observed in sensorimotor regions of interest in our paradigm is consistent with that observed during the bridging task. Our previous validation studies using electromyography (EMG) demonstrate that the rectus femoris muscle is activated to produce the focal movement for both the ULR and SLR.^22,23^ During the ULR, this is accompanied by an anticipatory postural activation synergy in the ipsilateral paraspinal and the bilateral abdominal muscles, and in the contralateral hip extensors. This postural activation helps to maintain the position of the spine and pelvis in the sagittal and transverse planes, in part by generating vertical ground reaction force under the contralateral foot.^4,22^ The leg raise is performed in a supine position, and therefore the mechanical conditions are different from those encountered during upright movement. However, APAs in the trunk musculature are preserved during limb movements even when the focal movement is performed in a less challenging context to postural equilibrium.^49^ In addition, de-Lima Pardini et al.,^4^ demonstrated that anticipatory ground reaction forces generated under the contralateral limb during the unsupported leg raise were significantly correlated with anticipatory ground reaction forces generated during gait initiation in standing. This suggests that postural control quantified using the fMRI leg raise paradigm is generalizable to that occurring during functional, upright voluntary movement.

In the two leg raise tasks used in this study, the goal is the same and the focal movements for both tasks are very similar. Therefore, as expected, both conditions elicited brain activation in regions associated with execution of a voluntary, goal-directed lower limb movement. The primary motor and somatosensory cortices contralateral to the moving limb were significantly activated during both the ULR and the SLR. The medial localization of this activation was consistent with the known somatotopic organization of both cortices.^50,51^ Both tasks also elicited activation within the premotor cortices. The activation within the premotor area in this study occurred medially and thus localizes to the SMA. The SMA is often loosely divided into pre-SMA and SMA proper, although these regions likely form a continuum.^52^ The area of activation evident in this study is consistent with SMA proper^53^, which is involved in the genesis and execution of movements that are self-initiated.^52,54^

During the ULR and SLR there was also activation in the primary somatosensory cortex. This is consistent with findings from previous studies investigating proprioceptive control of the trunk/proximal limbs.^38,51^ During movement, proprioceptive input from joint receptors and muscle spindles, as well as tactile information, is used to monitor movement and refine it as needed. This proprioceptive feedback is processed in Areas 2 and 3a of the contralateral primary somatosensory cortex.^25^ The sensory activation in both tasks also extended posteriorly into the contralateral medial superior parietal lobule. This region is believed to interpret sensory information as it relates to monitoring complex body positions within space and interpretation of spatial change during movement.^25,52^

We observed greater activation during the ULR than the SLR, in multiple cortical regions. Magnitude of activation was greater during the ULR in the contralateral (right) premotor cortex/SMA and in the ipsilateral (left) primary motor and somatosensory cortices, mid-cingulate cortex, and sensory association areas. Similarly, the extent of activation during the ULR was larger in the ipsilateral primary motor cortex, and lateralization of the primary somatosensory cortex was less. Increased activation in the left primary motor cortex was likely associated with the right-sided postural abdominal and hamstring activation that we have previously described during the ULR.^22,23^ Our findings confirm that primary motor cortex is involved in generating APAs in the contralateral trunk and hip musculature.^22,49^ The additional activation that we observed in the right SMA during the ULR was likely associated with the APA occurring in the left abdominal muscles during this task. Research using EEG or non-invasive brain stimulation has suggested that the SMA is involved with the timing^5,55^ and amplitude of APAs^14,20,56^ The SMA influences APAs directly via the corticospinal tract and indirectly as part of the cortico-basal ganglia-thalamo-cortical loop^56^ and the cortico-pontine-thalamo-cortical loop.^55^ We have previously demonstrated representation of the abdominal musculature within the somatotopic organization of SMA proper.^50^ In the same study we demonstrated that the representation of the abdominal musculature within SMA has greater functional connectivity with the putamen and cerebellum than the representation of the same musculature in M1. This supports the SMA’s role in shaping trunk muscle APAs via indirect loops.

Bilateral activation was evident in the supramarginal gyrus and parietal operculum during both tasks but was of significantly higher magnitude in the left hemisphere during the ULR. The border between these two regions is inconsistently defined and terminology varies across studies.^42,57^ Given this inconsistency, the ROI in our study encompassed both the supramarginal gyrus and the parietal operculum (Brodmann areas 40 and 43). Activation in the parietal operculum associates with attention to tactile stimuli, proprioception, and with processing of the sensory experience^25,58,59^ whereas activation in the supramarginal gyrus associates with proprioception.^42^ The significant activation in the left hemisphere during our active ULR contrasts with a previous study that indicated right laterality for proprioceptive activation in the supramarginal gyrus.^42^ However, this previous study involved proprioceptive activation occurring during passive rather than active movement of the right and left upper limbs.^42^ In our study, the increased activation in the left hemisphere during the ULR compared with the SLR was likely due to the interpretation and spatial processing of proprioceptive and tactile information associated with the postural muscle activation in the right trunk and lower limb.

During the ULR, compared to SLR, we also observed greater magnitude and extent of activation in the cerebellar vermis. Non-human primate studies suggest that projections from motor cortex to the cerebellar vermis are predominantly from the medial region of M1. Therefore, the vermis may be particularly involved in control of trunk/proximal musculature.^60^ In the cerebellar hemispheres, we focused on lobules III, IV, V, and VIII. These lobules receive input from sensorimotor cortical areas, particularly the premotor and primary motor cortices.^61^ Our findings of greater activation during the ULR are consistent with studies of individuals with cerebellar dysfunction indicating that the anterior cerebellum contributes to the timing and adaptability of APAs and other postural responses that occur during practiced movements.^18,62^ We also observed significant activation in the putamen bilaterally during the ULR but not during the SLR. Involvement of the putamen in anticipatory postural adjustments has also been demonstrated during focal movements involving the upper limbs in healthy adults,^20^ and impairments in anticipatory postural adjustments are evident in individuals with putamen dysfunction such as Parkinson’s disease.^63^ These findings suggest potential subcortical therapeutic targets for conditions characterized by APA abnormalities.

We acknowledge some limitations to this research. Given the poor temporal resolution of the BOLD hemodynamic response, the brain activation that we observed was not specific to the 150 ms window of time around the initiation of the focal movement that is considered an anticipatory postural adjustment.^1^ However, our paradigm did isolate the activation associated with the preparation to move following the Ready cue, and the initiation of the task following the Go cue. Additionally, some task comparisons for the ROI analysis did not reach significance following correction for multiple comparisons. It is probable that with greater study power we would have demonstrated significantly greater activation during the ULR in additional right hemispheric and sub-cortical ROIs. We also acknowledge the limitations of the analyses using the direct subtraction approach (contrast ULR – SLR). As has been noted for cognitive tasks, the relationship between the addition of components to a task and the representation of the more complex task in the brain is not purely additive or linear.^64^ For this reason we used multiple threshold and non-threshold dependent metrics, in addition to direct subtraction, for task comparisons. Despite the challenges of having participants move a lower limb within the scanner, we found that the framewise head displacement during the task was generally within acceptable limits following sufficient training and stabilization. In addition, the relaxation periods embedded within our event-related design ensured that multiple repetitions of the task were completed without discomfort or fatigue. Lastly, although our study population included only healthy adults, thus limiting generalizability to other populations, earlier preliminary work has also demonstrated the feasibility of this approach in older adults and adults with neurological dysfunction.^4^

## Conclusion

This study identified the brain activation associated with postural trunk and hip muscle control during voluntary lower limb tasks. Cortical and sub-cortical regions activated during the unsupported leg raise were consistent with the planning, execution, and sensory experience^65^ of a task involving multi-segmental and bilateral postural control, including anticipatory postural adjustments. This paradigm provides novel insights into sensorimotor events underlying leg raising and also serves as a foundation for future studies that will isolate neural mechanisms of impaired postural control in patients with neurological and musculoskeletal dysfunction.

## Data Availability

All data produced in the present study are available upon reasonable request to the authors

